# Evaluating Vaccine Efficacy Against SARS-CoV-2 Infection

**DOI:** 10.1101/2021.04.16.21255614

**Authors:** Dan-Yu Lin, Yu Gu, Donglin Zeng, Holly E. Janes, Peter B. Gilbert

## Abstract

Although interim results from several large placebo-controlled phase 3 trials demonstrated high vaccine efficacy (VE) against symptomatic COVID-19, it is unknown how effective the vaccines are in preventing people from becoming asymptomatically infected and potentially spreading the virus unwittingly. It is more difficult to evaluate VE against SARS-CoV-2 infection than against symptomatic COVID-19 because infection is not observed directly but rather is known to occur between two antibody or RT-PCR tests. Additional challenges arise as community transmission changes over time and as participants are vaccinated on different dates because of staggered enrollment or crossover before the end of the study. Here, we provide valid and efficient statistical methods for estimating potentially waning VE against SARS-CoV-2 infection with blood or nasal samples under time-varying community transmission, staggered enrollment, and blinded or unblinded crossover. We demonstrate the usefulness of the proposed methods through numerical studies mimicking the BNT162b2 phase 3 trial and the Prevent COVID U study. In addition, we assess how crossover and the frequency of diagnostic tests affect the precision of VE estimates.

**Summary:** We show how to estimate potentially waning efficacy of COVID-19 vaccines against SARS-CoV-2 infection using blood or nasal samples collected periodically from clinical trials with staggered enrollment of participants and crossover of placebo recipients.

## Introduction

Enormous progress has been made in the development of vaccines against severe acute respiratory syndrome coronavirus 2 (SARS-CoV-2). Within 1 year after the emergence of this novel infection that caused a global pandemic, vaccine targets were identified, vaccine constructs were created, and phase 1 through phase 3 testing was conducted. Interim results from several large-scale phase 3 randomized, placebo-controlled clinical trials have demonstrated high vaccine efficacy (VE) against symptomatic COVID-19 [1-4]. However, very little is known about VE against possibly asymptomatic SARS-CoV-2 infection.

It is critically important to assess VE against SARS-CoV-2 infection because reducing infection and community transmission is the key to halting the pandemic. Fortunately, most phase 3 trials have collected blood samples that can be used to identify SARS-CoV-2 sero-conversion [1-4]. For economic and logistical reasons, however, blood samples can only be drawn infrequently, such that seroconversion is only known to occur between two clinic visits that are weeks or months apart. It is more difficult to analyze such interval-censored seroconversion data than potentially right-censored symptomatic disease data, especially when VE changes over time. (An event time is said to be interval-censored if it is only known to lie in a time interval; an event time is said to be potentially right-censored if it is either observed exactly or known to be longer than the duration of follow-up [5].) Additional challenges arise when community transmission varies over time and when participants are vaccinated on different dates because of either staggered enrollment of participants or crossover of placebo participants to the vaccine arm before the end of the trial.

SARS-CoV-2 infection is commonly diagnosed by reverse transcription polymerase chain reaction (RT-PCR) on nasal swabs. Most phase 3 trials have collected nasal swabs at the enrollment and crossover visits [1-2, 4]. However, such infrequent swab samples will miss many infections, because a person may be RT-PCR positive for only a few days or weeks after infection [6]. Some phase 3 trials have taken nasal swabs more frequently (e.g., twice a week) on a subset of participants, and the newly launched Prevent COVID U study takes nasal swabs every day; however, frequent RT-PCR testing increases trial cost. How does the RT-PCR testing schedule affect the estimation of VE against infection (defined as viral RNA above a minimum threshold)?

In this article, we show how to evaluate potentially waning VE against SARS-CoV-2 infection — defined by seroconversion or detectable viral RNA — using blood or nasal samples taken at varying levels of frequency under the conditions of time-varying community transmission, staggered enrollment of participants, and possible crossover of placebo volunteers to the vaccine arm before the end of the study. We demonstrate the usefulness of the proposed methods through extensive simulation studies mimicking the BNT162b2 phase 3 trial [1] and the Prevent COVID U study. In addition, we investigate how the frequency of diagnostic tests and the characteristics (blinded versus unblinded, priority-dependent versus priority-independent) of crossover affect the precision of VE estimation.

## Methods

Figure 1 shows the blood sampling schedules for several phase 3 vaccine trials. For the three vaccines that have received FDA’s Emergency Use Authorization (EUA), blood samples are also taken at the crossover visits [1-2,4].

**Figure 1.**
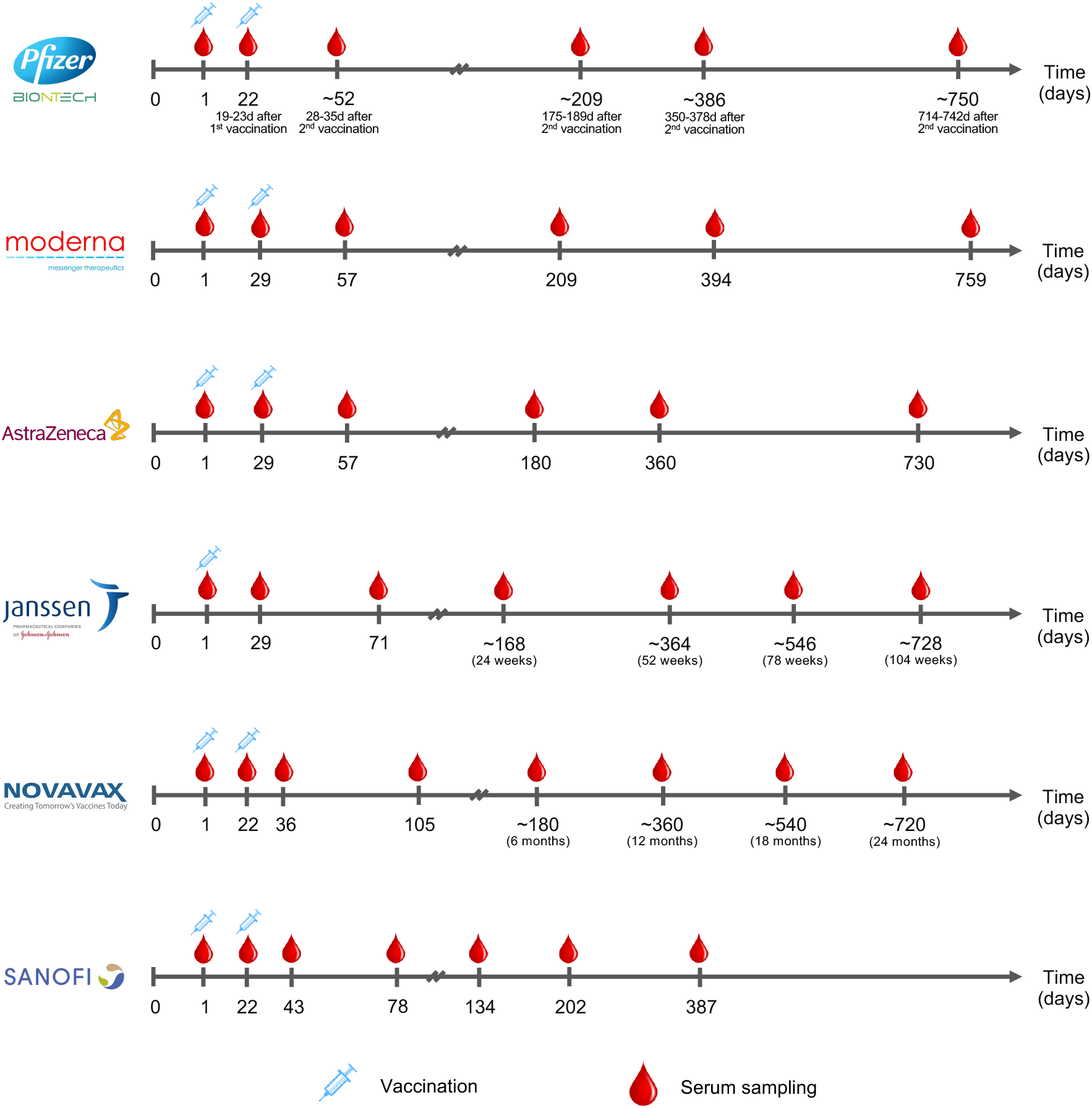
Serum sampling schedules in 6 phase 3 COVID-19 vaccine trials. The sampling time points are measured from the day of enrollment.

We are interested in time to SARS-CoV-2 infection assessed by seroconversion, which is only known to occur between two blood draws and is thus interval-censored. We allow the risk of infection to vary over the calendar time and to depend on baseline risk factors, such as age, sex, ethnicity, race, occupation, and health conditions; we allow the effect of vaccine on infection to depend on the time elapsed since vaccination.

We consider three measures of VE as a function of time elapsed since vaccination: (1) VE_h_(t) is the percentage reduction in the hazard rate or instantaneous risk of infection at time *t*; (2) VE_a_(t) is the percentage reduction in the attack rate or cumulative incidence of infection over the time period (0, *t*]; and (3) VE_a_(t_1_, t_2_) is the percentage reduction in the attack rate over the time period (*t*_1_, *t*_2_]. Note that VE_h_(t) and VE_a_(t) pertain to instantaneous and cumulative vaccine effects, respectively, and that VE_a_(t) is a special case of VE_a_(t_1_, t_2_) with *t*_1_ = 0 and *t*_2_ = *t*. If the vaccine effect is constant over time, then the three VE measures are equivalent (provided that the infection rate is low) [7].

In Supplementary Appendix 1, we formulate the above ideas through an adaptation of the well-known Cox [8] regression model, in which each participant’s time to infection is measured from the start of the clinical trial, and the hazard ratio of infection for vaccine versus placebo depends on the time elapsed since vaccination. Because of staggered enrollment and staggered crossover, the serum sampling time points are scattered randomly over time, providing valuable information about the distribution of the infection time. We express VE_h_ and VE_a_ as appropriate functions of the time-varying hazard ratio. We derive the maximum likelihood estimator for the time-varying hazard ratio based on the interval-censored infection time data and provide the corresponding estimators of VE_h_ and VE_a_.

The above framework also applies to RT-PCR tests of nasal swabs. Because an infected person is RT-PCR positive for a shorter period of time than they are seropositive (days/weeks versus months) [6,9-10], nasal swabbing needs to be done more frequently than serum sampling in order to capture the infections defined by detectable viral RNA. With very frequent RT-PCR, time to detectable viral RNA may be treated as a potentially right-censored event time. In our framework, potentially right-censored data is a special case of interval-censored data, with the exactly observed event time lying within an interval of one day.

## Results

We conducted a series of simulation studies mimicking the BNT162b2 phase 3 trial [1] (Supplementary Appendix 2.1). We used a total of 40,000 participants who enter the trial at a constant rate over a 4-month period and are randomly assigned to vaccine or placebo in a 1:1 ratio. To reflect the increase of COVID-19 cases since last summer and the downward trend this spring, we let the risk of infection increase over the first 7 months and decrease afterward. In addition, we let the risk of infection depend strongly on priority tier.

As in the BNT162b2 phase 3 trial [1], the vaccine in our simulation received an EUA from FDA at the 5th month, after which placebo participants are sequentially crossed over to the vaccine arm. We considered:

### Priority-dependent crossover

Crossover starts at month 6, 7, 8, 9, or 10 of the study for participants with priority tier of 1, 2, 3, 4, or 5, respectively, each participant’s waiting time for the clinic visit following the exponential distribution with mean of 0.5 month.

### Priority-independent crossover

Crossover starts at month 6 of the study for all participants, with the waiting time following the exponential distribution with mean of 0.5 month.

Note that crossover spreads over a longer time period under priority-dependent crossover than under priority-independent crossover.

We considered both blinded and unblinded crossover. At blinded crossover, placebo participants receive the vaccine and vaccine participants receive the placebo; none of them are aware of the order of their treatments. At unblinded crossover, participants are notified of their original treatment assignments, and placebo recipients are vaccinated. In both types of crossover, all participants are followed until the time of analysis, which is 10.5 months since trial initiation. To avoid bias due to behavioral confounding, we discarded the data collected after unblinded crossover.

As shown in Figure 1, blood samples were scheduled to be drawn on Day 1, Day 22, Day 52 and Day 209 (during the first year) in the BNT162b2 phase 3 trial [1]. In our simulation, we allowed for small random deviations from the schedule. Blood samples were also drawn at the crossover visits. Because of staggered enrollment and staggered crossover, serum sampling points were scattered randomly over the study period, making it possible to estimate time-varying VE.

We also simulated a design under which there is no crossover before the time of analysis. Without crossover, placebo participants stay on placebo longer than with crossover, providing more information about long-term placebo-controlled VE. However, because crossover is one of the serum sampling points, there are fewer sampling points and thus fewer antibody tests under no crossover than under crossover.

Naturally, VE_h_ equals 0 at the first injection. We let VE_h_ increase from 0 to 80% at 4 weeks and then either stay constant or decrease gradually over time. We refer to these two situations as constant VE and waning VE, respectively. (Note that constant versus waning VE pertains only to the period after the first 4 weeks, when VE is ramping up.)

In our first simulation scenario, we let VE_h_ stay at 80% after 4 weeks, and we analyzed the resulting interval-censored data using the proposed method with a log hazard ratio that decreases linearly between weeks 0 and 4 and stays constant after week 4. For comparison, we fit the same model by treating the time of the first positive antibody test as a potentially right-censored event time and performing maximum partial likelihood estimation [5,8]; we refer to this approach as naive Cox regression. (Naive Cox regression estimates the same VE parameter using the same data as the proposed method, the only difference being that it converts interval-censored event times to potentially right-censored event times.) For further comparison, we implemented logistic regression by treating the seroconversion status at the last blood test before the 10.5 month mark (excluding the blood samples drawn after unblinded crossover) as a binary outcome and estimating the odds ratio of seroconversion between the vaccine and the placebo groups by the maximum likelihood estimator.

Table 1 summarizes the results of these simulation studies. Using the proposed method, the VE estimates are unbiased, the standard errors are accurately estimated, and the confidence intervals have proper coverage probabilities. The standard error is lower under blinded than unblinded crossover, and lower under priority-dependent than priority-independent crossover. The standard error is slightly higher under no crossover than under blinded priority-dependent crossover. (Note that there are fewer sampling points under no crossover than under crossover.) In comparison, naive Cox regression may under-estimate or over-estimate the true VE, and logistic regression always under-estimates the true VE.

**Table 1.** **Estimation of Constant VE Based on Antibody Tests Under No Crossover (A), Blinded Priority-Dependent (B) and Priority-Independent (C) Crossover, and Unblinded Priority-Dependent (B’) and Priority-Independent (C’) Crossover When** VE_h_ **Stays at 80% After Week 4**

| Design | Proposed Method |  |  |  | Naive Cox Regression |  |  |  | Logistic Regression |  |  |  |
| --- | --- | --- | --- | --- | --- | --- | --- | --- | --- | --- | --- | --- |
|  | Mean | SE | SEE | CP | Mean | SE | SEE | CP | Mean | SE | SEE | CP |
| A | 79.8% | 1.33% | 1.33% | 94% | 76.8% | 1.42% | 1.43% | 33% | 77.1% | 1.37% | 1.38% | 39% |
| B | 79.9% | 1.31% | 1.30% | 95% | 87.3% | 0.95% | 0.97% | 0% | 75.2% | 1.47% | 1.46% | 5% |
| C | 79.9% | 1.58% | 1.54% | 93% | 89.6% | 1.13% | 1.12% | 0% | 70.6% | 1.83% | 1.80% | 0% |
| B' | 79.8% | 1.41% | 1.38% | 95% | 76.9% | 1.49% | 1.47% | 40% | 76.9% | 1.45% | 1.43% | 37% |
| C' | 79.8% | 1.67% | 1.63% | 95% | 75.8% | 1.80% | 1.78% | 27% | 75.4% | 1.76% | 1.74% | 17% |

Note: Mean and SE denote the mean and standard error of the VE estimator, SEE denotes the mean of the standard error estimator, and CP denotes the coverage probability of the 95% confidence interval.

In our second simulation scenario, we let VE_h_ stay at 80% after 4 weeks or let it decrease to 0 at 1 year. We implemented the proposed method (for interval-censored data) using a piecewise linear function for the log hazard ratio, with a change point placed at 4 weeks and with the two slopes estimated from the data. For comparison, we also implemented naive Cox regression with the same piecewise linear function for the log hazard ratio.

Tables 2–3 show the simulation results on the estimation of VE_a_ over successive time periods under constant VE and waning VE, respectively. The proposed method yields unbiased VE_a_ estimates, with accurate standard error estimates and proper confidence intervals in virtually all cases. Naive Cox regression yields severely biased VE_a_ estimates.

**Table 2.** **Estimation of** VE_a_ **Over Successive Time Periods Based on Antibody Tests Under No Crossover (A), Blinded Priority-Dependent (B) and Priority-Independent (C) Crossover, and Unblinded Priority-Dependent (B’) and Priority-Independent (C’) Crossover When VE Does Not Wane Over Time**

| Design | Weeks | True | Proposed Method |  |  |  | Naive Cox Regression |  |  |  |
| --- | --- | --- | --- | --- | --- | --- | --- | --- | --- | --- |
| | | $VE_a$ | Mean | SE | SEE | CP | Mean | SE | SEE | CP |
| A | 0-4 | 50.3% | 49.9% | 3.2% | 3.0% | 94% | 48.5% | 4.0% | 3.9% | 92% |
|  | 4-16 | 80.0% | 79.7% | 2.2% | 2.1% | 94% | 77.6% | 3.3% | 3.2% | 86% |
|  | 16-28 | 80.0% | 79.9% | 1.9% | 1.8% | 94% | 76.9% | 1.4% | 1.4% | 36% |
|  | 28-40 | 80.0% | 79.7% | 4.6% | 4.2% | 92% | 75.8% | 3.4% | 3.3% | 74% |
| B | 0-4 | 50.3% | 50.1% | 2.2% | 2.2% | 94% | 59.9% | 1.7% | 1.7% | 0% |
|  | 4-16 | 80.0% | 79.8% | 1.7% | 1.6% | 93% | 88.5% | 1.2% | 1.2% | 0% |
|  | 16-28 | 80.0% | 80.0% | 1.6% | 1.6% | 95% | 87.0% | 1.0% | 1.0% | 0% |
|  | 28-40 | 80.0% | 80.0% | 3.1% | 3.0% | 95% | 85.3% | 1.5% | 1.5% | 14% |
| C | 0-4 | 50.3% | 50.1% | 2.1% | 2.0% | 94% | 61.1% | 1.8% | 1.7% | 0% |
|  | 4-16 | 80.0% | 79.9% | 1.7% | 1.7% | 94% | 89.9% | 1.2% | 1.2% | 0% |
|  | 16-28 | 80.0% | 80.0% | 2.0% | 2.0% | 95% | 89.3% | 1.2% | 1.2% | 0% |
|  | 28-40 | 80.0% | 79.9% | 3.6% | 3.5% | 95% | 88.7% | 1.7% | 1.7% | 2% |
| B' | 0-4 | 50.3% | 49.7% | 2.6% | 2.5% | 95% | 39.6% | 3.2% | 3.4% | 6% |
|  | 4-16 | 80.0% | 79.6% | 1.8% | 1.8% | 95% | 70.8% | 2.8% | 2.9% | 5% |
|  | 16-28 | 80.0% | 80.1% | 1.9% | 1.8% | 94% | 77.3% | 1.5% | 1.5% | 51% |
|  | 28-40 | 80.0% | 80.2% | 3.9% | 3.8% | 94% | 82.3% | 2.1% | 2.1% | 82% |
| C' | 0-4 | 50.3% | 49.6% | 2.9% | 2.8% | 94% | 37.8% | 3.9% | 3.9% | 7% |
|  | 4-16 | 80.0% | 79.6% | 1.8% | 1.8% | 94% | 70.8% | 2.9% | 2.9% | 4% |
|  | 16-28 | 80.0% | 80.2% | 3.3% | 3.2% | 95% | 80.2% | 2.2% | 2.2% | 96% |
|  | 28-40 | 80.0% | 80.1% | 6.9% | 6.5% | 94% | 86.2% | 3.4% | 3.3% | 62% |

Note: See Note to Table 1.

**Table 3.** **Estimation of** VE_a_ **Over Successive Time Periods Based on Antibody Tests Under No Crossover (A), Blinded Priority-Dependent (B) and Priority-Independent (C) Crossover, and Unblinded Priority-Dependent (B’) and Priority-Independent (C’) Crossover When VE Wanes Over Time**

| Design | Weeks | True | Proposed Method |  |  |  | Naive Cox Regression |  |  |  |
| --- | --- | --- | --- | --- | --- | --- | --- | --- | --- | --- |
| | | $VE_a$ | Mean | SE | SEE | CP | Mean | SE | SEE | CP |
| A | 0-4 | 50.3% | 50.3% | 3.1% | 2.7% | 91% | 58.3% | 3.6% | 3.5% | 42% |
|  | 4-16 | 75.3% | 75.3% | 2.6% | 2.3% | 92% | 82.9% | 3.0% | 2.9% | 39% |
|  | 16-28 | 62.9% | 62.9% | 2.7% | 2.9% | 96% | 68.0% | 1.9% | 1.9% | 27% |
|  | 28-40 | 44.3% | 43.2% | 10.8% | 10.3% | 93% | 38.6% | 8.4% | 8.2% | 91% |
| B | 0-4 | 50.3% | 50.1% | 2.1% | 2.0% | 95% | 61.4% | 1.6% | 1.5% | 0% |
|  | 4-16 | 75.3% | 75.2% | 1.9% | 1.9% | 94% | 87.6% | 1.2% | 1.2% | 0% |
|  | 16-28 | 62.9% | 63.1% | 2.3% | 2.3% | 96% | 79.8% | 1.3% | 1.3% | 0% |
|  | 28-40 | 44.3% | 44.7% | 6.6% | 6.5% | 94% | 66.8% | 2.8% | 2.8% | 0% |
| C | 0-4 | 50.3% | 50.2% | 1.9% | 1.9% | 94% | 62.5% | 1.6% | 1.6% | 0% |
|  | 4-16 | 75.3% | 75.3% | 1.9% | 1.9% | 95% | 88.8% | 1.2% | 1.3% | 0% |
|  | 16-28 | 62.9% | 63.0% | 3.1% | 3.0% | 95% | 82.1% | 1.7% | 1.7% | 0% |
|  | 28-40 | 44.3% | 44.1% | 8.0% | 7.9% | 95% | 71.4% | 3.6% | 3.6% | 0% |
| B' | 0-4 | 50.3% | 49.9% | 2.3% | 2.3% | 95% | 43.5% | 2.7% | 2.9% | 28% |
|  | 4-16 | 75.3% | 75.0% | 2.0% | 2.0% | 96% | 70.2% | 2.7% | 2.8% | 50% |
|  | 16-28 | 62.9% | 62.9% | 2.6% | 2.5% | 95% | 66.3% | 1.8% | 1.9% | 56% |
|  | 28-40 | 44.3% | 44.5% | 8.1% | 7.9% | 95% | 61.8% | 3.5% | 3.5% | 1% |
| C' | 0-4 | 50.3% | 49.7% | 2.6% | 2.6% | 95% | 41.7% | 3.4% | 3.4% | 22% |
|  | 4-16 | 75.3% | 74.9% | 2.0% | 2.1% | 95% | 69.3% | 2.9% | 2.9% | 37% |
|  | 16-28 | 62.9% | 63.1% | 4.8% | 4.8% | 94% | 68.4% | 2.7% | 2.8% | 56% |
|  | 28-40 | 44.3% | 44.5% | 15.1% | 14.8% | 94% | 67.0% | 6.4% | 6.5% | 19% |

Note: See the Note to Table 1.

Figure 2 displays the estimation results produced by the proposed method in one of the trials simulated under waning VE. The estimated VE_h_ and VE_a_ curves are close to the truth, and the 95% confidence intervals nearly cover the entire true curves. As expected, the confidence intervals are the narrowest under blinded priority-dependent crossover and the widest under unblinded priority-independent crossover. In addition, the confidence intervals for VE_h_ are wider than the confidence intervals for VE_a_ at the right tail.

**Figure 2.**
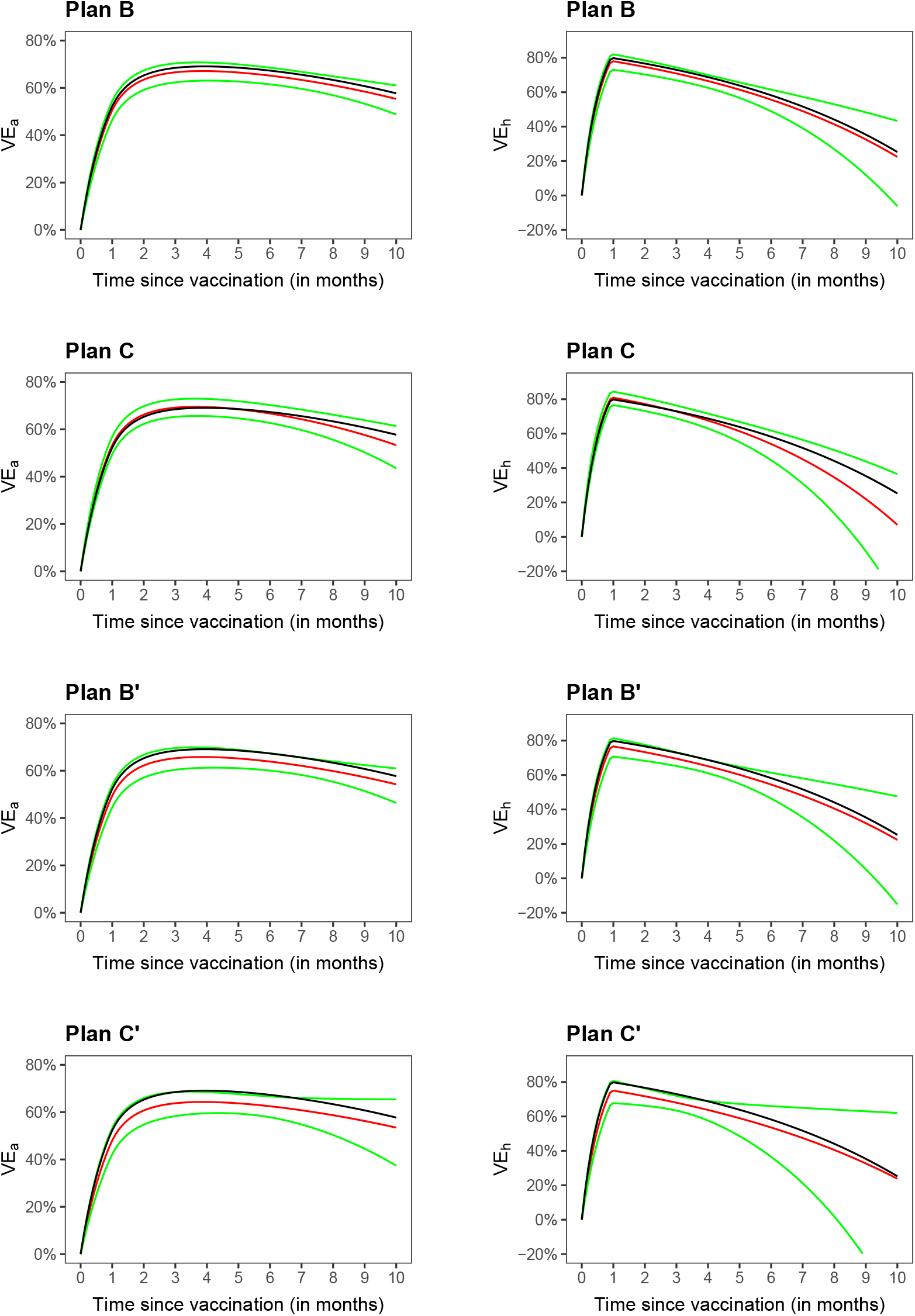
Estimation of VE_a_ and VE_h_ in a clinical trial under blinded priority-dependent (B) and priority-independent (C) crossover and under unblinded priority-dependent (B’) and priority-independent (C’) crossover: the black curve pertains to the true value, the red curve to the proposed estimate, and the green curves to the 95% confidence intervals.

We also conducted a series of simulation studies mimicking the Prevent COVID U study (Supplementary Appendix 2.2). A total of 12,000 participants enter the study at a constant rate over one month. Half of them are randomly selected to receive the Moderna vaccine at enrollment, and the other half get their first injection with a 4 month delay. We assumed a downward trend of infection over time; we adopted the VE patterns from the first series of simulation studies but placed the change point at 6 weeks instead of 4 weeks.

We explored various swabbing/RT-PCR testing schedules, ranging from every day to every 2 weeks. Each participant is followed for 4 months, and the study ends at month 5, when the last enrolled participant has been followed for 4 months. We also considered a scenario where 50% of the delayed-arm participants receive outside vaccines before the end of follow-up; in the analysis, we discarded the data collected after outside vaccination.

Table 4 summarizes the simulation results on the estimation of constant VE. The proposed method yields unbiased VE estimates, with accurate standard error estimates and proper confidence intervals in all cases. The standard error of the VE estimate tends to increase a little bit as RT-PCR testing becomes less frequent. There is a slight loss of precision in the VE estimates when the delayed-arm participants are allowed to receive outside vaccines. In comparison, naive Cox regression shows highly similar results to the proposed method when RT-PCR testing is performed every day; however, as RT-PCR testing becomes less frequent, naive Cox regression becomes more biased, with increasingly larger standard error than the proposed method. Excluding the events within the first 6 weeks substantially reduces the precision of VE estimates. (The substantial loss of precision is due to relatively high incidence in the first 6 weeks under decreasing background incidence over time and high VE. In the setting of constant background incidence with VE of 0.6 after 4 weeks, excluding the events within the first 4 weeks incurs about 13% loss of statistical efficiency.)

**Table 4.** **Estimation of Constant VE Under Different RT-PCR Testing Schedules When** VE_h_ **Stays at 80% After Week 6**

| Outside<br>Vaccines | Frequency<br>of Tests | Proposed Method |  |  |  |  | Naive Cox Regression |  |  |  |
| --- | --- | --- | --- | --- | --- | --- | --- | --- | --- | --- |
|  |  | Mean | SE | SEE | CP | RE | Mean | SE | SEE | CP |
| No | Every day | 79.5% | 3.44% | 3.49% | 95% | 1.36 | 79.5% | 3.44% | 3.46% | 95% |
|  | Every 2 days | 79.4% | 3.52% | 3.51% | 95% | 1.35 | 79.1% | 3.54% | 3.50% | 94% |
|  | Every 4 days | 79.5% | 3.49% | 3.52% | 95% | 1.38 | 78.7% | 3.53% | 3.53% | 93% |
|  | Every week | 79.5% | 3.50% | 3.49% | 95% | 1.33 | 77.7% | 3.59% | 3.55% | 90% |
|  | Every 2 weeks | 79.5% | 3.58% | 3.60% | 95% | 1.35 | 75.4% | 3.81% | 3.80% | 74% |
| Yes | Every day | 79.4% | 3.50% | 3.52% | 95% | 1.35 | 79.4% | 3.50% | 3.49% | 95% |
|  | Every 2 days | 79.5% | 3.48% | 3.53% | 95% | 1.36 | 79.2% | 3.49% | 3.51% | 95% |
|  | Every 4 days | 79.5% | 3.58% | 3.56% | 95% | 1.35 | 78.7% | 3.60% | 3.56% | 93% |
|  | Every week | 79.5% | 3.53% | 3.52% | 95% | 1.34 | 77.9% | 3.59% | 3.56% | 91% |
|  | Every 2 weeks | 79.4% | 3.69% | 3.64% | 95% | 1.35 | 75.7% | 3.85% | 3.79% | 76% |

Note: Mean and SE denote the mean and standard error of the VE estimator, SEE denotes the mean of the standard error estimator, CP denotes the coverage probability of the 95% confidence interval, and RE is the variance of the VE estimator excluding the infections of the first 6 weeks divided by the variance of the VE estimator using all infections.

Table 5 presents the results on the estimation of VE_a_ over successive time periods when VE_h_ decreases to 0 at 1 year. The proposed method provides unbiased VE_a_ estimates, along with proper confidence intervals. Naive Cox regression performs well when RT-PCR testing is done daily but performs poorly when RT-PCR testing is infrequent.

**Table 5.**
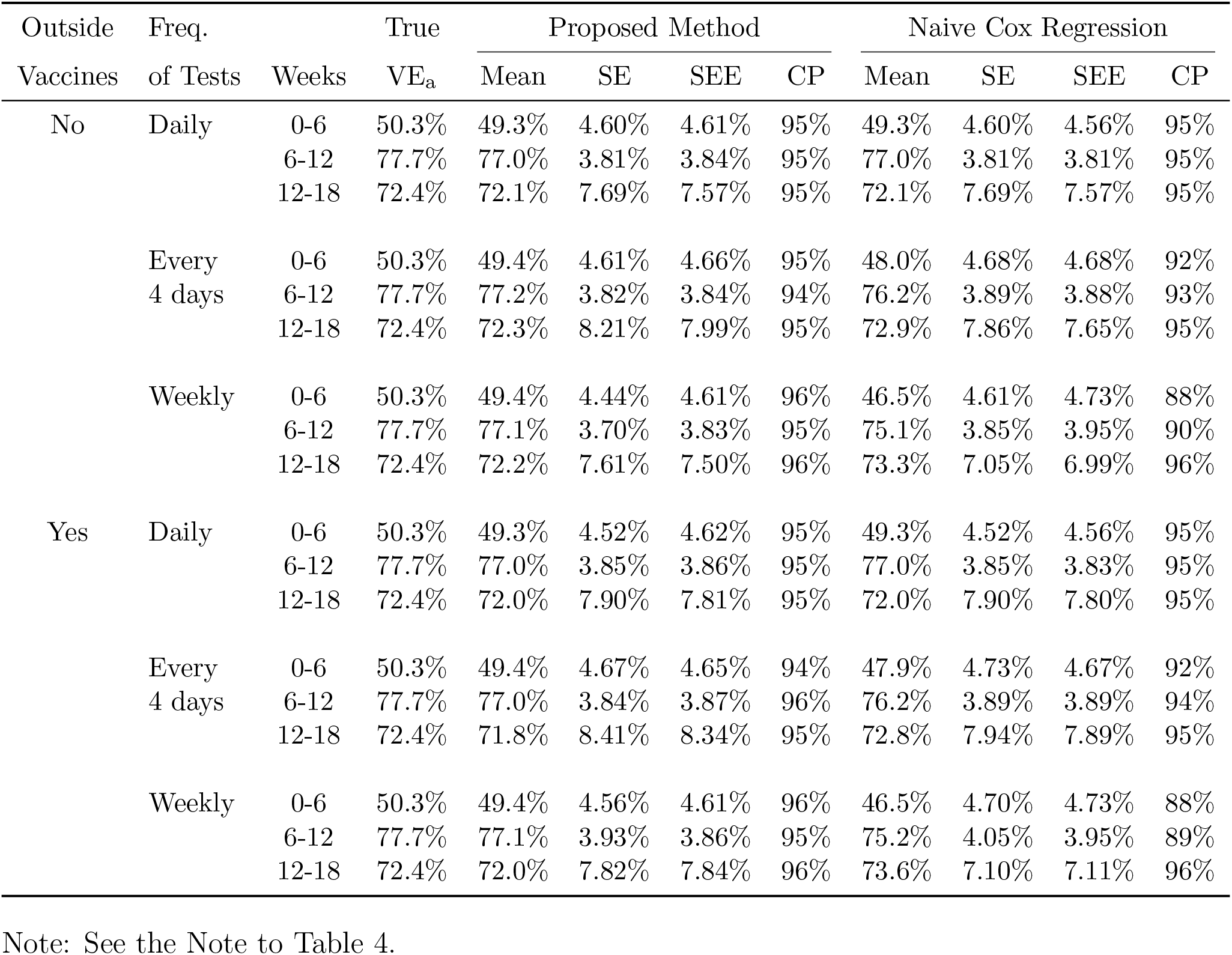
**Estimation of** VE_a_ **Over Successive Time Periods Under Different RT-PCR Testing Schedules When VE Wanes Over Time**

We have assumed that VE ramps to an unknown peak level 4 weeks (or 6 weeks) after the first injection of the Pfizer/BioNTech vaccine (or the Moderna vaccine). We can allow uncertainty in this change point by including several change points in the analysis or by selecting the change point through the Akaike information criterion (AIC). We evaluated these strategies by extending the simulation studies reported in Table 1. We considered two scenarios: (a) the true change point is 4 weeks; and (b) the true change point is 6 weeks. In both scenarios, the true VE increases from 0 at time 0 to 0.8 at the true change point and stays at 0.8 afterward. We implemented two methods: (1) place 3 change points at weeks 4, 6, and 8 and estimate the corresponding 3 slopes of the log hazard ratio; and (2) calculate the likelihood with the change point placed at week 4, 6, or 8 and select the time point that yields the highest likelihood. As shown in Table 6, the first method performs very well, although the standard error is higher than using a single change point. The second method correctly selects the change point with high probability.

**Table 6.** **Estimation of Constant VE With Unknown Change Points Under No Crossover (A), Blinded Priority-Dependent (B) and Priority-Independent (C) Crossover, and Unblinded Priority-Dependent (B’) and Priority-Independent (C’) Crossover When** VE_h_ **Stays at 80% After the Change Point**

| Design | (a) True change point = 4 weeks |  |  |  |  | (b) True change point = 6 weeks |  |  |  |  |
| --- | --- | --- | --- | --- | --- | --- | --- | --- | --- | --- |
|  | Mean | SE | SEE | CP | Correct | Mean | SE | SEE | CP | Correct |
| A | 79.9% | 1.50% | 1.47% | 94% | 73% | 79.9% | 1.46% | 1.47% | 94% | 65% |
| B | 80.0% | 1.42% | 1.42% | 95% | 80% | 79.9% | 1.43% | 1.42% | 95% | 73% |
| C | 79.9% | 1.78% | 1.70% | 94% | 82% | 79.9% | 1.73% | 1.69% | 95% | 78% |
| B' | 79.9% | 1.58% | 1.52% | 94% | 72% | 79.9% | 1.60% | 1.53% | 94% | 67% |
| C' | 79.9% | 1.96% | 1.90% | 94% | 70% | 79.9% | 1.91% | 1.91% | 95% | 67% |

Note: Mean and SE denote the mean and standard error of the VE estimator, SEE denotes the mean of the standard error estimator, and CP denotes the coverage probability of the 95% confidence interval, when VE may reach its peak level at one of three change points. Correct denotes the probability of correctly selecting the change point by the AIC.

## Discussion

We have demonstrated that it is possible to evaluate time-varying VE against SARS-CoV-2 infection using the blood samples collected in the ongoing phase 3 vaccine trials [1-4] or using the nasal samples collected in studies like Prevent COVID U. We found that when antibody or RT-PCR tests are performed infrequently, the use of standard Cox regression for potentially right-censored data yields biased and imprecise VE estimates. The new methods provide valid and efficient estimation of three useful VE measures.

The form of the model considered in this article is essentially the same as that used in our previous work on evaluating VE against symptomatic COVID-19 [7]. However, here the estimation approach is different because infection times are interval-censored rather than potentially right-censored. The proposed methodology is general enough to include potentially right-censored data as a special case and thus offers an alternative way to assess VE against symptomatic COVID-19. A major advantage of this new approach is that it provides a unified framework for studying constant versus waning VE.

Another important contribution of this work is a careful treatment of the ramping VE after initial vaccination. The prevailing approach is not to count the events that occur within 4–6 weeks of the first injection [1-2]. Discarding the first 4–6 weeks of follow-up data causes considerable loss of statistical efficiency, as shown in Table 4. In the case of blinded crossover, excluding the events that occur within 4–6 weeks of crossover will further reduce statistical efficiency, whereas including all the events will result in biased VE estimates.

We have not accounted for the measurement errors of antibody or RT-PCR tests in the analysis. The false-positive rate is negligible for RT-PCR testing and small for antibody testing. An infected person is seropositive for a longer period of time than they are RT-PCR positive (several months versus several days or weeks) [6, 9-10]. Thus, infrequent serology will capture more infections than infrequent RT-PCR. Some asymptomatic infections never seroconvert or have transient seroconversion that may be missed by infrequent serology [9-10]. However, those who do not seroconvert tend to be less infectious than those who do, such that missed seronegative infections may be clinically less important. Likewise, an asymptomatic infection that is RT-PCR positive for just a day or two is difficult to detect but may have little public-health relevance.

For the Prevent COVID U study, the main reason for daily swabbing and testing is not to determine the timing of infection but rather to measure the full course of viral load for all infected participants. In particular, investigators wish to capture potential infectiousness by measuring the peak viral load, the duration of viral shedding, and the area under the viral load curve. If detecting the presence of viral RNA were the study’s only goal, then less frequent testing would be needed. The proposed methods (for interval-censored data) may be warranted in the case that a substantial number of swabs are not collected or are not usable (due to improper collection or storage).

Blood samples and nasal swabs provide complementary information about SARS-CoV-2 infection. Viral RNA can be detected sooner after infection than seroconversion, but antibody lasts longer than viral shedding [6, 9-10]. There is considerable heterogeneity in the duration of both seropositivity and RT-PCR positivity, with the biggest driving factor being symptomatic versus asymptomatic infection [6, 9-10]. Many studies collect both blood and nasal samples. For example, the Moderna phase 3 trial [2] performs RT-PCR testing at month 1 and at crossover in addition periodic serology. The Prevent COVID U study performs periodic (every 2 months) serology and frequent RT-PCR testing. The proposed methods can be applied to the two types of infection data separately or as a combined endpoint, depending on the objective of the analysis and the frequency of each type of test.

The monitoring times are assumed to be independent of the infection time (conditional on covariates). This assumption is satisfied for planned diagnostic tests but is unlikely to hold if SARS-CoV-2 infection is detected through symptom-prompted testing. We can apply the proposed methods (for interval-censored data) to planned tests and standard Cox regression with potentially right-censored data to symptom-prompted tests. If the planned RT-PCR testing is frequent, then the data from planned and symptom-prompted RT-PCR tests can be combined and standard Cox regression for potentially right-censored data can be adopted.

We have implemented the methods described in this article in an R package, which is available at https://dlin.web.unc.edu/software/idove/.

## Data Availability

N/A

https://dlin.web.unc.edu/software/idove/

## Authors’ Information

Dan-Yu Lin, Ph.D., is Dennis Gillings Distinguished Professor of Biostatistics, Yu Gu is doctoral student, and Donglin Zeng, Ph.D., is Professor of Biostatistics, Gillings School of Global Public Health, University of North Carolina, Chapel Hill, NC 27599-7420, USA. Holly E. Janes, Ph.D., and Peter B. Gilbert, Ph.D., are Professors, Vaccine and Infectious Disease Division, Fred Hutch, Seattle, WA 98109-1024, USA.

## Acknowledgments

This work was supported by the NIH grants R01 AI029168, R01 GM124104, P01 CA142538, and UM1 AI068635.

## Supplementary Appendix

### 1. Statistical Methods

Let *S* denote the time when the participant is vaccinated, and *T* denote the time when the participant acquires SARS-CoV-2 infection (as defined by seroconversion or detectable viral RNA), with both times measured in days from the start of the clinical trial. In addition, let *X* denote baseline risk factors (e.g., age, occupation, race, health conditions). We specify that the hazard function of *T* is related to *S* and *X* through the Cox [1] regression model

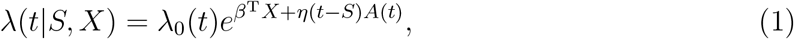

where *A*(*t*) = *I*(*S < t*), *I*(·) is the indicator function, λ_0_(·) is an arbitrary baseline hazard function, *β* is a set of regression parameters representing the effects of baseline risk factors, and *η*(·) is the log hazard ratio characterizing the time-varying effect of vaccination. Under this formulation, the baseline hazard function varies over the calendar time, and the effect of vaccine on the risk of infection depends on the time elapsed since vaccination.

We can define the vaccine efficacy at day *t* as the proportionate reduction in the hazard rate of infection at day *t* for individuals who were vaccinated *t* days ago compared with those who have not been vaccinated, i.e., *V E*_*h*_(*t*) = 1 *− e*^*η*(*t*)^. In addition, we can define the *t*-day vaccine efficacy as the proportionate reduction in the attack rate or cumulative incidence of infection by day *t* for individuals who were vaccinated *t* days ago compared with the non-vaccinated individuals:

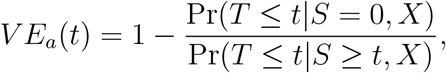

which is approximately 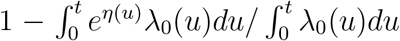 when the infection rate is low. If λ_0_(·) is approximately constant, then *V E*_*a*_(*t*) = 1 *− V* (*t*)*/t*, where 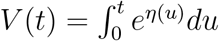. Finally, we consider the vaccine efficacy in reducing the attack rate over a certain time period, say (*t*_1_, *t*_2_]:

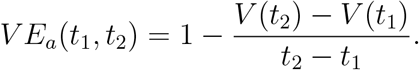

Clearly, all three VE measures are simple functions of the log hazard ratio *η*(·).

For economic and logistical reasons, antibody (or RT-PCR) tests can only be performed infrequently for each participant. Thus, SARS-CoV-2 infection, as defined by seroconversion or detectable viral RNA, is only known to occur over a broad time interval, such that *T* must be treated as an interval-censored event time. Let *L* be the time of the last negative test, and *R* be the time of the first positive test, such that *T* is known to lie in the time interval (*L, R*]. (To be more precise, *L* is the last seronegative test date minus 7 days and *R* is the first seropositive test date minus 7 days, because it takes approximately 7 days after initial SARS-CoV-2 acquisition for the antibody test to register positive.) In addition, let *E* be the time when the participant enters the clinical trial. Like *T* and *S*, the time variables *E, L*, and *R* are measured from the start of the clinical trial. We assume that *E, L, R*, and *S* are independent of *T* conditional on *X*.

For a clinical trial with a total of *n* participants, the data consist of (*E*_*i*_, *L*_*i*_, *R*_*i*_, *S*_*i*_, *X*_*i*_) (*i* = 1,…, *n*). The likelihood takes the form

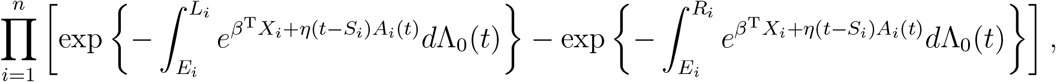

where 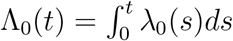.

This likelihood involves two infinite-dimensional functions Λ_0_(·) and *η*(·), which are not identifiable if both are unrestricted. We let Λ_0_(·) be completely nonparametric and estimate it by a step function with non-negative jumps at the unique values of *L*_*i*_ *>* 0 and *R*_*i*_ *< ∞* (*i* = 1,…, *n*). We approximate *η*(·) by a sequence of B-splines functions, denoted by *B*_1_(*t*),…, *B*_*K*_(*t*), such that 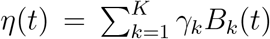. Write *γ* = (*γ*_1_,…, *γ*_*K*_)^T^, and *Z*_*i*_(*t*) = (*B*_1_(*t − S*_*i*_)*A*_*i*_(*t*),…, *B*_*K*_(*t − S*_*i*_)*A*_*i*_(*t*))^T^. Then the likelihood becomes

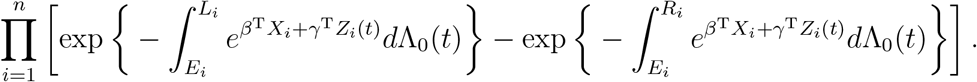

This is the interval-censored data likelihood for the standard Cox model with time-independent covariates *X* and time-dependent covariates *Z* [2], except that the event time is measured from the start of the study rather than the participant’s entry time. We compute the nonparametric maximum likelihood estimator for (*β, γ*, Λ_0_), denoted by 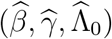, through an EM algorithm based on latent Poisson random variables [2]. We then estimate *η*(*t*) and *V* (*t*) by 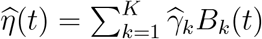 and 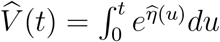, respectively.

By appealing to modern empirical process theory [3], we can show that 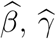, and 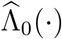 are consistent. In addition, 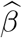 and 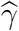 are asymptotically normal and their covariance matrix can be consistently estimated by the Hessian matrix [2,4] of the profile log-likelihood for (*β, γ*), where the log-likelihood is maximized with respect to Λ_0_ for fixed *β* and *γ* via the EM algorithm. These results allow us to estimate *V E*_*h*_(*t*), *V E*_*a*_(*t*), and *V E*_*a*_(*t*_1_, *t*_2_), construct confidence intervals, and perform hypothesis testing.

#### Remark

In our previous work on (potentially right-censored) symptomatic disease, we approximate log λ_0_(·) by *B*-spline functions while letting *η*(·) be completely nonparametric [5]. With interval-censored data, a completely nonparametric function cannot be estimated at the parametric rate, making it difficult to construct confidence intervals. Thus, we let λ_0_(·) be completely nonparametric and approximate *η*(·), which is the parameter of main interest, by *B*-spline functions. One benefit of this approach is that it provides a unified framework to study constant versus time-varying VE (by choosing appropriate *B*-spline functions). This framework also unifies the analysis of symptomatic disease and asymptomatic infection because potentially right-censored data can be treated as a special case of interval-censored data. For potentially right-censored data, we can adopt very flexible *B*-spline functions for *η*(·); for truly interval-censored data, we have to be more rigid unless the sample size is very large, the infection rate is high, or antibody/RT-PCR tests are performed frequently.

### 2. Simulation Studies

#### 2.1. Antibody Tests

We designed the first series of simulation studies to mimic the BNT162b2 phase 3 trial [6]. We assumed that 40,000 participants entered the study at a constant rate over four months, i.e., *E ∼* Uniform(0, 4) months. (In the actual trial, the number of participants was slightly below 40,000, after exclusion of those who were seropositive at baseline.) We created a composite baseline risk score *X*, which takes values 1, 2, 3, 4, and 5 with equal probability. We randomly assigned half of the participants at study entry to vaccine and half to placebo.We generated the infection time *T* from model (1) with *β* = 0.2 and

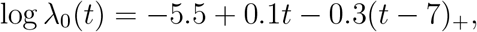

where *t*_+_ = *t* if *t >* 0 and 0 otherwise. We assumed that *V E*_*h*_ starts at 0 at *t* = 0, increases to some maximum value at *t* = *t*_*m*_, and then stays constant or decreases gradually over time. Specifically, we set *η*(*t*) = *b*_1_*t* (0 *≤ t < t*_*m*_) and *η*(*t*) = *b*_1_*t*_*m*_ + *b*_2_(*t − t*_*m*_) (*t ≥ t*_*m*_), where *t*_*m*_ = 4 weeks, and *b*_1_ and *b*_2_ were chosen such that *V E*_*h*_(*t*_*m*_) = 0.8 and *V E*_*h*_(1 year) = 0.8 or 0.

In the BNT162b2 phase 3 trial [6], serum samples were scheduled to be drawn on Day 1, Day 22, Day 52, and Day 209 (as measured from the participant’s entry time) [6]. To allow for small random departures from the schedule, we used Day 1, Day 22 + Uniform (–1,3) days, Day 52 + Uniform(–2, 8) days, and Day 209 + Uniform(–5,10) days.

Serum samples were also drawn at the crossover visits. We considered:

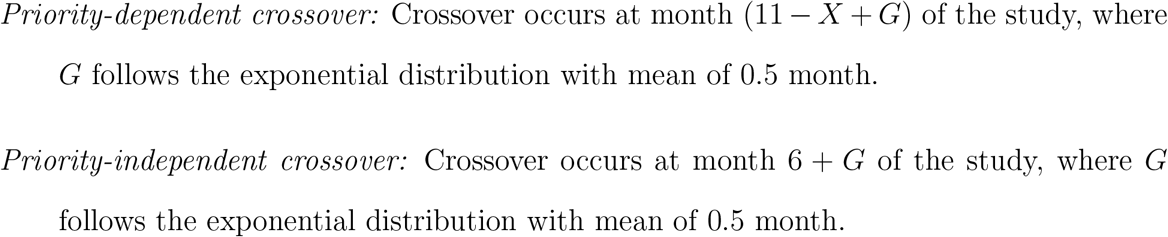

We assumed that the analysis is performed at 10.5 months after the start of the study, such that only the blood samples that were drawn before the 10.5 month mark can be included. We considered both blinded and unblinded crossover designs. Under blinded crossover, participants receive the opposite of their original assignments, and we used all the data that are collected before the time of analysis. At the point of unblinded crossover, participants are notified of their original assignments, and placebo participants receive the vaccine; we disregarded any data collected after unblinded crossover in order to avoid bias due to behavioral confounding.

We applied the proposed methods to each simulated dataset by setting *η*(*t*) in model (1) to be piecewise linear with a change point placed at *t*_*m*_ and with the slope after *t*_*m*_ fixed at 0 or estimated from data. For comparison, we performed maximum partial likelihood estimation of the same model with the same data by treating the time of the first positive antibody test as a potentially right-censored event time. We also performed the logistic regression of the infection status at the last antibody test before the 10.5 month mark (excluding the blood samples drawn after unblinded crossover) on the randomization assignment and baseline covariates *X*, and we estimated VE by 1 minus the odds ratio of infection for vaccine versus placebo.

#### 2.2. RT-PCR Tests

We conducted a second series of simulation studies to mimic the Prevent COVID U study. In our simulation, a total of 12,000 participants enter the study at a constant rate over one month; half of them receive the Moderna vaccine at enrollment and the other half 4 months later. We generated the infection time *T* from model (1) without *X* and with log λ_0_(*t*) = *−*4.0 *−* 0.2*t*. We assumed the same VE patterns as in the first series of simulation studies, but the change point was set at 6 weeks instead of 4 weeks.

We investigated various swabbing/RT-PCR testing schedules, ranging from every day to every 2 weeks. All participants are followed for 4 months, and the study ends at month 5. In addition, we considered a scenario in which placebo participants may receive vaccines outside of the study 1 month after enrollment. We assumed that the time to outside vaccination follows the Weibull distribution with shape parameter of 3 and scale parameter of 4, such that the cumulative probability of outside vaccination is approximately 50%.

For each simulated dataset, we analyzed the data in the same way as in the first series of simulation studies. Specifically, we implemented both the proposed method and its right-censored data counterpart, to be referred to as naive Cox regression. We discarded the data collected on placebo participants after they received outside vaccines.

